# Risky Sexual Practices and Associated Factors among Taxi and Three-Wheeled Vehicle Drivers in Arba Minch Town, Southern Ethiopia: A Community-Based Cross-Sectional Study

**DOI:** 10.64898/2026.05.01.26352270

**Authors:** Fintew Eshetu, Tesfaye Feleke, Gebremaryam Temesgen, Negusse Boti Sidamo

## Abstract

**Background:** Risky sexual practices contribute significantly to the transmission of sexually transmitted infections (STIs) and HIV/AIDS. Taxi and three-wheeled vehicle drivers may be particularly vulnerable due to occupational mobility and related lifestyle factors. However, evidence on this population in Ethiopia remains limited.

**Methods:** A community-based cross-sectional study was conducted among taxi and three-wheeled vehicle drivers in Arba Minch town, Ethiopia, from July 26 to August 26, 2025. Participants were selected using a simple random sampling technique. Data were collected using an interviewer-administered questionnaire through the Kobo Toolbox mobile application. Data were exported to SPSS version 27 for cleaning and analysis. Binary logistic regression analysis was performed to identify factors associated with current risky sexual practices. Adjusted odds ratios (AORs) with 95% confidence intervals (CIs) were used, and statistical significance was declared at p < 0.05.

**Results:** A total of 640 drivers participated in the study (response rate: 97.6%). The prevalence of lifetime risky sexual practices was 46.9% (95% CI: 43.0%–50.8%), while the prevalence of current risky sexual practices was 37.2% (95% CI: 33.4%–41.1%). Factors independently associated with current risky sexual practices included living alone (AOR = 3.01; 95% CI: 2.03–4.45), not discussing sexual and reproductive health issues (AOR = 2.09; 95% CI: 1.17–3.75), substance use (AOR = 1.69; 95% CI: 1.10–2.61), attending nightclubs (AOR = 2.04; 95% CI: 1.41–2.96), and exposure to pornographic materials (AOR = 2.19; 95% CI: 1.49–3.23).

**Conclusion:** A substantial proportion of taxi and three-wheeled vehicle drivers reported engaging in risky sexual practices, indicating a significant and overlooked public health concern within this occupational group. These practices were independently associated with modifiable behavioral and lifestyle factors, including substance use, nightlife attendance, and inadequate communication on sexual and reproductive health issues. The findings underscore the urgent need for innovative, multi-sectoral interventions that extend beyond conventional health service delivery. Integrating basic sexual and reproductive health education and risk-reduction messaging into driver licensing and refresher training programs may provide a feasible and scalable opportunity to reach this mobile and high-risk population. Such integration, in collaboration with transport authorities and public health sectors, has the potential to improve awareness, promote safer behaviors, and reduce vulnerability to risky sexual practices among drivers.

## Background

Risky Sexual Practices refer to behaviors that increase the likelihood of adverse Sexual and Reproductive Health (SRH) outcomes, including multiple sexual partnerships, inconsistent condom use, and early sexual initiation. These behaviors are key contributors to the transmission of sexually transmitted infections (STIs), including HIV, and to unintended pregnancies, particularly in low- and middle-income countries (1).

Globally, STIs remain a major public health challenge, with disproportionate burden in sub-Saharan Africa(2). Behavioral, structural, and occupational factors contribute to persistent engagement in risky sexual practices among mobile and hard-to-reach populations. Among these groups, transport workers, particularly taxi and three-wheeled vehicle drivers, have been identified as potentially vulnerable due to their occupational mobility, prolonged working hours, frequent night travel, and extended periods away from regular sexual partners (3).

Evidence suggests that such occupational conditions may increase exposure to high-risk environments, including nightlife settings and transient sexual partnerships (4). In addition, limited access to preventive services, such as condoms and SRH information during work hours, may further exacerbate vulnerability. Individual-level factors such as substance use, including alcohol, khat, and other psychoactive substances, have also been consistently associated with impaired sexual decision-making and increased engagement in risky sexual behaviors(5). Furthermore, peer influence and social norms, as well as inadequate communication about SRH issues within families and social networks, have been shown to shape sexual behaviors in this population(6).

Studies conducted in different settings, including Ethiopia, have reported a high prevalence of risky sexual practices among transport workers, with factors such as substance use, nightlife exposure, and low condom utilization repeatedly identified as key correlates (7, 8). However, existing evidence remains fragmented and largely concentrated in urban centers outside southern Ethiopia, with limited context-specific data on taxi and three-wheeled vehicle drivers in Arba Minch town (9, 10).

Despite ongoing national efforts to improve sexual and reproductive health outcomes and reduce the burden of HIV/AIDS and STIs, risky sexual behaviors continue to persist among high-risk occupational groups(11–13)(14). (15). Understanding the behavioral and contextual determinants of these practices among drivers is essential for designing targeted, evidence-based interventions.

Therefore, this study aimed to assess risky sexual practices and associated factors among taxi and three-wheeled vehicle drivers in Arba Minch town, Southern Ethiopia.

## Methods and Materials

### Study Setting and Period

This study was conducted in the *Gamo zone*, which is located in the southern regional state of Ethiopia. Arba Minch Town is located approximately 505 km south of Addis Ababa and has an estimated population of 201,049 (101,030 males and 100,019 females), based on projections from the 2022 Central Statistical Agency census (16). The town is administratively divided into eight sub-cities (Secha, Bere-Edgetber, Gurba, Nechsar, Sikela, Lemat, Bola-Gurba, and Shara-Chano), further subdivided into 24 kebeles.

According to the Arba Minch Town Transport Office (2023), the town has a registered fleet of 3,299 vehicles, including taxis and three-wheeled vehicles, which are primarily used for public transportation and constitute the focus of this study due to their occupational mobility and associated behavioral risk exposure.

Participant recruitment started on 26/07/2025 and ended on 26/08/2025.

### Study design

A community-based cross-sectional study was conducted in Arba Minch Town, Southern Ethiopia

### Source and Study Population

The source population comprised all taxi and three-wheeled vehicle drivers operating in Arba Minch Town during the study period. The study population consisted of selected eligible taxi and three-wheeled vehicle drivers who met the inclusion criteria and were available during data collection.

### Eligibility Criteria

Drivers were eligible for inclusion if they had worked as taxi or three-wheeled vehicle operators for at least six months prior to data collection and were aged 18 years or older. Drivers who were severely ill, unable to communicate, or unwilling to provide informed consent at the time of data collection were excluded from the study.

### Sample Size Determination

The sample size was calculated by using two population proportion formulas using EPI Info software version 7. Using Factors associated with risky sexual practice, like multiple sexual partners,premarital sex, Use of condoms during sex and condom use status (inconsistent and consistent). Taking a confidence interval 95%, power =80% and a ratio of (unexposed: exposed) 1 to 1. The final sample size was 656.

### Sampling Procedure

A stratified simple random sampling technique was employed. The study population was stratified into two groups: taxi drivers and three-wheeled vehicle drivers. The sampling frame was obtained from official registration lists of driver associations in Arba Minch Town, which include one taxi association and seven three-wheeled vehicle associations. Unique identification numbers from association logbooks were used to construct the sampling frame. Proportional allocation was applied to determine the sample size for each stratum based on population size. Accordingly, 18 participants were selected from 60 taxi drivers, and 638 participants were selected from 2,100 three-wheeled vehicle drivers. Within each stratum, participants were selected using a computer-generated simple random sampling method based on their unique identification numbers.

### Operational Definition

#### Risky sexual practices

Engagement in one or more of the following behaviors: having multiple sexual partners, sexual intercourse with casual partners, sexual intercourse with commercial sex workers, or inconsistent condom use (5, 17).

### Lifetime risky sexual practices

Drivers who have one or more of the following sexual practices: having multiple sexual partners, having sexual intercourse with a casual sexual partner or having sexual intercourse with a commercial sex worker, or inconsistent use of condoms in the lifetime (5, 17).

### Current risky sexual practices

Drivers who have one or more of the following sexual practices: having multiple sexual partners, having sexual intercourse with a casual sexual partner or having sexual intercourse with a commercial sex worker, or inconsistent use of condoms in the past 6 months (5, 17).

### Substance use

Alcohol consumption, khat chewing, smoking, use of shisha and related substances that alter the individual’s conscious judgment of the individual (18).

### Sexually Active

Drivers who had at least one sexual encounter before the survey(19).

### Inconsistent condom use

Interrupted, incorrect, and occasional use of a condom during episodes of sexual activity with at least one non-married and extra sexual partner(20).

### Knowledge about SRH issue

Assessed using 15-item questionnaire; correct responses were scored 1 and incorrect responses 0. Participants scoring ≥75% of the total score were categorized as having sufficient knowledge, while those scoring <75% were considered to have insufficient knowledge.

### Attitude about SHR issue

Measured using 8 Likert-scale items. A composite score was computed, and participants scoring ≥75% were classified as having a favorable attitude, while those scoring below 75% were classified as having an unfavorable attitude.

### Data collection Procedure

A pre-tested, structured questionnaire was used to collect data through face-to-face interviews. The data collection tool was developed after a thorough review of previously published articles to ensure its relevance and alignment with established research standards (41, 49).To ensure the quality of data,the questionnaire was first prepared in English and translated into Amharic and retranslated back to English to check for consistency.To ensure the quality and consistency of data collection, daily meetings were held involving the principal investigator and data collectors. These meetings provided a platform to discuss progress, address challenges, and ensure the smooth functioning of the data collection process.A pre-test was conducted on 33(5%) of the sample size, 2 weeks before the actual data collection period in Birbir town, and necessary adjustments were made.

### Data Processing and Analysis

Data were collected electronically using Kobo Toolbox and exported to IBM SPSS Statistics version 27 for analysis. Prior to statistical analysis, the dataset was assessed for completeness, consistency, and plausibility. Data cleaning procedures included range and logic checks, recoding of variables, and handling of missing values based on predefined criteria.

The internal consistency of composite measures (knowledge and attitude) was evaluated using Cronbach’s alpha, with values of 0.83 and 0.81, respectively, indicating good reliability.

Composite scores were computed by summing item responses, and where applicable, categorized using predefined or literature-supported cut-off points.

Descriptive statistics were used to summarize study variables in accordance with STROBE recommendations. Continuous variables were assessed for normality and summarized using means and standard deviations (or medians and interquartile ranges where appropriate), while categorical variables were presented as frequencies and percentages.

To identify factors associated with risky sexual practices, binary logistic regression analysis was performed. Initially, bivariable logistic regression analyses were conducted, and variables with a p-value ≤ 0.25 were considered candidates for the multivariable model to minimize residual confounding. A multivariable logistic regression model was then fitted to estimate independent associations while controlling for potential confounders. Model adequacy was assessed using the Hosmer–Lemeshow test (p = 0.798), indicating good model fit. Multicollinearity among independent variables was evaluated using the variance inflation factor (VIF), with acceptable thresholds applied. Adjusted odds ratios (AORs) with 95% confidence intervals (CIs) were calculated to quantify the strength and direction of associations. Statistical significance was declared at a two-sided p-value < 0.05. All analyses adhered to standard epidemiological reporting practices, and results were presented using text, tables, and figures to ensure clarity and transparency.

## Ethical Considerations

Ethical clearance was obtained from the Institutional Review Board (IRB) of Arba Minch University, College of Medicine and Health Sciences (Ref. No. IRB/23338/25). Following approval, an official letter of cooperation was issued by the School of Public Health to the relevant authorities. Prior to data collection, appropriate communication was made with responsible bodies to explain the purpose and procedures of the study. Written informed consent was obtained from all participants after clearly explaining the study’s objectives, potential benefits, and possible risks. Participants were assured of their right to refuse participation or withdraw from the study at any stage without any consequences. Participation was entirely voluntary, and respondents were given the opportunity to ask questions before and during the interview. Confidentiality and privacy were strictly maintained throughout the study. No personal identifiers were recorded, and all information collected was used solely for research purposes. Participants were assured that their responses would not be disclosed to anyone outside the research team. All study procedures were conducted in accordance with the principles of the Declaration of Helsinki.

## Result

### Socio-demographic characteristics of study participants

A total of 640 participants were included in the study, yielding a response rate of 640 (97.6%). The mean age of participants was 26.47 (SD ± 5.33) years. The majority, 270 (42.2%), were aged 25–34 years, followed by 267 (41.7%) aged 18–24 years. More than half of the participants, 326 (51.0%), were followers of the Orthodox religion, and 330 (51.6%) were unmarried. Regarding living arrangements, 374 (58.4%) lived with their families, while 266 (41.6%) lived alone. Nearly half, 292 (45.6%), owned their vehicles. In terms of work experience, 339 (53.0%) had ≥5 years of experience. Additionally, 380 (59.4%) reported a monthly income of ≥5000 Ethiopian Birr **(Table 1).**

**Table 1:** Socio-demographic characteristics of taxi and three-wheeled drivers in Arba Minch town, south Ethiopia, 2025 (n = 640)

| Variables | Categories | Frequency(n) | Percentage (%) |
| --- | --- | --- | --- |
| **Age(year)** | 18 – 24 | 267 | 41.7 |
|  | 25 – 34 | 270 | 42.2 |
|  | ≥ 35 | 103 | 16.1 |
| **Religion** | Orthodox | 326 | 51 |
|  | Muslim | 128 | 20 |
|  | Protestant | 171 | 26.7 |
|  | Other* | 15 | 2.3 |
| **Ethnicity** | Gamo | 382 | 59.7 |
|  | Wolaita | 95 | 14.8 |
|  | Amhara | 133 | 20.8 |
|  | Konso | 30 | 4.7 |
| **Marital status** | Unmarried | 330 | 51.6 |
|  | Married | 279 | 43.6 |
|  | Others** | 31 | 4.8 |
| **Educational status** | Completed primary school | 100 | 15.6 |
|  | Completed secondary school | 288 | 45 |
|  | College or above | 252 | 39.4 |
| **Living arraignment** | Live alone | 266 | 41.6 |
|  | Live with family | 374 | 58.4 |
| **Vehicle ownership** | Owner | 292 | 45.6 |
|  | Not owner | 348 | 54.4 |
| **Work experience** | Less than 5 years | 301 | 47 |
|  | ≥ 5 years | 339 | 53 |
| **Monthly Income** | < 5000 ETB | 260 | 40.6 |
|  | ≥5,000 ETB | 380 | 59.4 |

\* Only Jesus and seventh day Adventist
\*\*Divorce and Widowed

### Behavioral characteristics of respondents

Among the participants, 328 (51.2%) reported substance use. Among substance users (n = 328), 115 (35.1%) reported khat chewing, 103 (31.4%) alcohol consumption, and 83 (25.3%) cigarette smoking.

More than half, 366 (57.2%), reported having intimate friends or partners who used substances, and 284 (44.4%) reported peer initiation of sexual activity. Regarding sexual and reproductive health (SRH) communication, 276 (43.1%) reported no discussion with family or peers.

Additionally, 251 (39.2%) reported watching pornography, and 258 (40.3%) reported attending nightclubs. A history of sexually transmitted infections in the past six months was reported by 293 (45.8%) participants **(Table 2).**

**Table 2:** Behavioral factors of taxi and three-wheel drivers in Arba Minch town, south Ethiopia, 2025.

| Variables | Categories | Frequency(n) | Percentage (%) |
| --- | --- | --- | --- |
| Substance use | Yes | 328 | 51.2 |
|  | No | 312 | 48.8 |
| Type of substance (n=328) | Alcohol | 103 | 31.4 |
|  | Khat | 115 | 35.1 |
|  | Cigarette | 83 | 25.3 |
|  | Other* | 20 | 6.1 |
|  | More than 2 type of substances | 7 | 2.1 |
| Frequency Alcohol use (n=103) | Every day | 22 | 21.3 |
|  | Every week | 25 | 24.3 |
|  | Sometimes | 31 | 30.1 |
|  | On Holyday | 25 | 24.3 |
| Frequency Khat use (n=115) | Every day | 24 | 20.9 |
|  | Every week | 36 | 31.3 |
|  | Sometimes | 33 | 28.7 |
|  | On Holyday | 22 | 19.1 |
| **Frequency Cigarette (n=83)** | Every day | 27 | 32.5 |
|  | Every week | 19 | 22.9 |
|  | Sometimes | 21 | 25.3 |
|  | On Holyday | 16 | 19.3 |
| **Intimate friends/partners use substances** | Yes | 366 | 57.2 |
|  | No | 274 | 42.8 |
| **Intimate friend who start sex** | Yes | 284 | 44.4 |
|  | No | 356 | 55.6 |
| **SRH issue discussion with family/ Peer** | No | 276 | 43.1 |
|  | Yes | 364 | 56.9 |
| **Watching pornography (n=640)** | Yes | 251 | 39.2 |
|  | No | 389 | 60.8 |
| **Attending night club (n=640)** | Yes | 258 | 40.3 |
|  | No | 382 | 59.7 |
| **History of STI (n=640)** | Yes | 293 | 45.8 |
|  | No | 347 | 54.2 |

\*Shisha and related illicit drugs

### Knowledge of sexual and reproductive health issues

Overall, 347 (54.2%) participants had sufficient knowledge of sexual and reproductive health issues. The mean knowledge score was 20.52 (SD ± 3.91). The majority, 514 (80.3%), had heard about STIs/HIV. Regarding transmission, 417 (65.2%) identified unprotected sexual intercourse, and 361 (56.4%) identified mother-to-child transmission. Regarding prevention, 530 (82.8%) reported consistent condom use as a preventive method. Knowledge of symptoms included genital itching, reported by 446 (69.7%), and genital discharge, reported by 442 (69.1%) **(Table 3).**

**Table 3:**
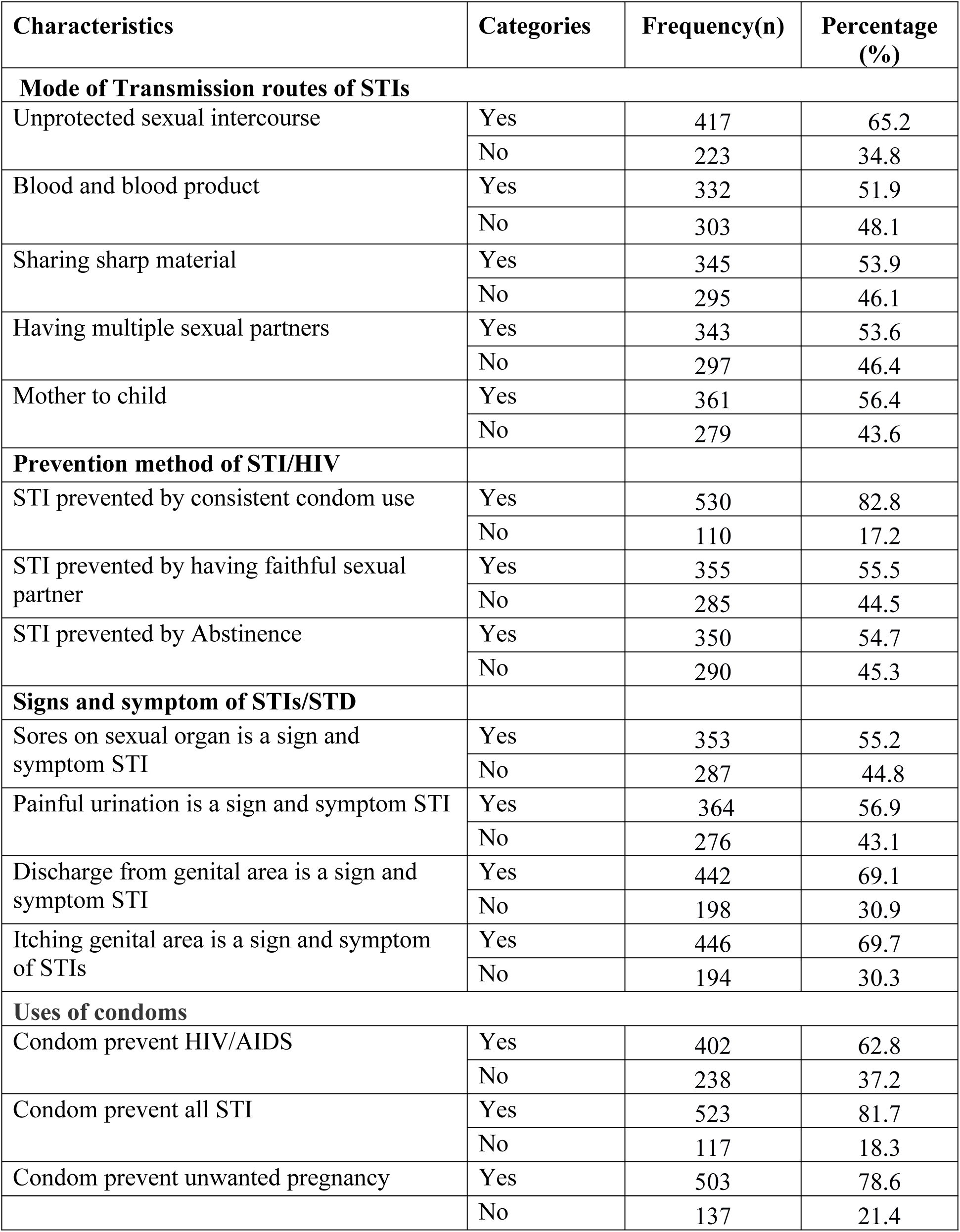
Knowledge about SRH of taxi and three-wheeled drivers in Arba Minch town, South Ethiopia, 2025.

| Characteristics | Categories | Frequency(n) | Percentage (%) |
| --- | --- | --- | --- |
| **Mode of Transmission routes of STIs** |  |  |  |
| Unprotected sexual intercourse | Yes | 417 | 65.2 |
|  | No | 223 | 34.8 |
| Blood and blood product | Yes | 332 | 51.9 |
|  | No | 303 | 48.1 |
| Sharing sharp material | Yes | 345 | 53.9 |
|  | No | 295 | 46.1 |
| Having multiple sexual partners | Yes | 343 | 53.6 |
|  | No | 297 | 46.4 |
| Mother to child | Yes | 361 | 56.4 |
|  | No | 279 | 43.6 |
| **Prevention method of STI/HIV** |  |  |  |
| STI prevented by consistent condom use | Yes | 530 | 82.8 |
|  | No | 110 | 17.2 |
| STI prevented by having faithful sexual partner | Yes | 355 | 55.5 |
|  | No | 285 | 44.5 |
| STI prevented by Abstinence | Yes | 350 | 54.7 |
|  | No | 290 | 45.3 |
| **Signs and symptom of STIs/STD** |  |  |  |
| Sores on sexual organ is a sign and symptom STI | Yes | 353 | 55.2 |
|  | No | 287 | 44.8 |
| Painful urination is a sign and symptom STI | Yes | 364 | 56.9 |
|  | No | 276 | 43.1 |
| Discharge from genital area is a sign and symptom STI | Yes | 442 | 69.1 |
|  | No | 198 | 30.9 |
| Itching genital area is a sign and symptom of STIs | Yes | 446 | 69.7 |
|  | No | 194 | 30.3 |
| **Uses of condoms** |  |  |  |
| Condom prevent HIV/AIDS | Yes | 402 | 62.8 |
|  | No | 238 | 37.2 |
| Condom prevent all STI | Yes | 523 | 81.7 |
|  | No | 117 | 18.3 |
| Condom prevent unwanted pregnancy | Yes | 503 | 78.6 |
|  | No | 137 | 21.4 |

### Attitudes toward sexual and reproductive health issues

A total of 351 (54.8%) participants had a favorable attitude toward sexual and reproductive health issues. The mean attitude score was 3.68 (SD ± 0.74). Half of the participants, 322 (50.3%), agreed that STIs can be prevented. Additionally, 391 (61.1%) agreed that having only one sexual partner is beneficial, and 345 (53.9%) agreed that abstinence reduces the risk of HIV infection **(Table 4).**

**Table 4:** Attitude toward Sexual and Reproductive Health issues among taxi and three-wheeled drivers in Arba Minch Town, South Ethiopia, 2025.

| Characteristics | Strongly disagree | Disagree | Not sure | Agree | Strongly agree |
| --- | --- | --- | --- | --- | --- |
| STI can be cured | 87 (13.6%) | 102(15.9%) | 194(30.3%) | 136(21.3%) | 123(19.2%) |
| STI can be prevented | 55 (8.6%) | 63(9.8%) | 50(7.8%) | 322(50.3%) | 150(23.4%) |
| Condom effectively protect against HIV | 53 (8.3%) | 45 (7.0%) | 49 (7.7%) | 348(54.4%) | 145(22.7%) |
| Condom effectively protect against pregnancy | 45 (7.0%) | 36 (5.6%) | 48 (7.5%) | 341(53.3%) | 170(26.6%) |
| Condom effectively protect against STI | 58 (9.1%) | 48 (7.5%) | 51 (8.0%) | 343(53.6%) | 140(21.9%) |
| Being abstain from sex is an effective means of reducing one's own Risk of HIV Infection | 51 (8.0%) | 68 (10.6%) | 63 (9.8%) | 345(53.9%) | 113(17.7%) |
| It would okay to have more than one sexual partner | 52 (8.1%) | 46 (7.2%) | 52 (8.1%) | 344(53.8%) | 146(22.8%) |
| Having only one partner is good for my partner as well as for me | 35 (5.5%) | 30 (4.7%) | 41 (6.4%) | 391(61.1%) | 143(22.3%) |

### Risky Sexual Practices among taxi and three-wheeled drivers

Among the 640 respondents, 340 (53.1%) reported ever having sexual intercourse. The age at first sexual intercourse ranged from 15 to 24 years, with a mean age of 17.83 years (SD = 2.26). A substantial proportion, 223 (65.6%), initiated sexual intercourse before the age of 18 years, and 177 (52.1%) reported that their first sexual experience occurred before joining their current occupation. Among sexually active participants (n = 340), several lifetime risky sexual behaviors were identified. Specifically, 102 (30.0%) reported having multiple sexual partners, 100 (29.4%) reported inconsistent condom use, and 105 (30.8%) reported ever having sexual intercourse with a commercial sex worker. Overall, 300 (46.9%) (95% CI: 43.0%–50.8%) of all respondents had engaged in at least one form of risky sexual practice during their lifetime. Regarding recent behaviors, 238 (37.2%) (95% CI: 33.4%–41.1%) respondents reported engaging in risky sexual practices within the past six months. Among these, 128 (53.8%) reported multiple sexual partners, 103 (43.3%) reported inconsistent condom use, and 138 (58.0%) reported sexual intercourse with commercial sex workers during the same period.

### Factors Associated with Risky Sexual Practices

Both bivariable and multivariable logistic regression analyses were conducted to identify factors associated with risky sexual practices among taxi and three-wheel drivers. In the bivariable analysis, variables including age, educational status, living arrangement, vehicle ownership status, peers initiating sexual activity, history of STIs, substance use, pornography viewing, nightclub attendance, discussion of SRH issues, and attitude towards SRH were associated with risky sexual practices at a p-value ≤ 0.25 and were therefore considered for the multivariable model.

In the multivariable logistic regression analysis, living arrangement, substance use, lack of SRH discussion, nightclub attendance, and pornography consumption remained statistically significant predictors of risky sexual practices at p < 0.05 with 95% confidence intervals.

Participants who lived alone had approximately three times higher odds of engaging in risky sexual practices compared with those living with family members (AOR = 3.01; 95% CI: 2.03–4.45). Similarly, respondents who did not discuss SRH issues had about twice the odds of risky sexual practices compared to those who engaged in such discussions (AOR = 2.09; 95% CI: 1.17–3.75).

Substance use was also significantly associated with risky sexual practices; substance users had 1.69 times higher odds compared to non-users (AOR = 1.69; 95% CI: 1.10–2.61). In addition, participants who attended nightclubs were twice as likely to engage in risky sexual practices compared to their counterparts (AOR = 2.04; 95% CI: 1.41–2.96). Likewise, those who reported watching pornography had higher odds of risky sexual practices than those who did not (AOR = 2.19; 95% CI: 1.49–3.23) **(Table 5).**

**Table 5:** Bivariable and multivariable logistic regression analysis of factors associated with risky sexual practices among taxi and three-wheeled vehicle drivers in Arba Minch, Southern Ethiopia, 2025.

| Variables | Categories: | Risky Sexual Practices |  | COR (95% CI) | AOR (95% CI) | P-value |
| --- | --- | --- | --- | --- | --- | --- |
|  |  | Risky | Non-Risky |  |  |  |
| Age | 18 – 24 | 82 | 185 | 0.82 (0.51–1.34) | 0.62 (0.35–1.08) | 0.09 |
|  | 25 – 34 | 120 | 150 | 1.48 (0.93–2.38) | 0.90 (0.53–1.55) | 0.70 |
|  | ≥ 35 | 36 | 67 | 1 (Ref) | 1 (Ref) | — |
| Educational status | Primary | 30 | 70 | 0.57 (0.34–0.93) | 0.59 (0.34–1.03) | 0.06 |
|  | Secondary | 100 | 188 | 0.70 (0.50–1.00) | 0.71 (0.48–1.05) | 0.09 |
|  | College or above | 108 | 144 | 1 (Ref) | 1 (Ref) | — |
| Living arrangement | Alone | 130 | 108 | 2.35(1.69-3.26) | 3.01(2.03-4.45) | **0.000\*\*** |
|  | With family | 136 | 266 | 1 | 1 |  |
| Vehicle owner | Owner | 117 | 121 | 1.25(0.90-1.73) | 1.27(0.83-1.96) | 0.2 |
|  | Non-owner | 175 | 227 | 1 | 1 |  |
| Peers start sex | Yes | 119 | 119 | 1.43(1.04-1.98) | 1.28(0.89-1.85) | 0.1 |
|  | No | 165 | 237 | 1 | 1 |  |
| Ever had STDs | Yes | 121 | 117 | 1.38(1.00-1.90) | 0.58(0.32-1.07) | 0.8 |
|  | No | 117 | 230 | 1 | 1 |  |
| Substance use | Yes | 138 | 100 | 1.54(1.11-2.12) | 1.69(1.10-2.61) | **0.016\*** |
|  | No | 190 | 212 | 1 | 1 |  |
| Watching pornography | Yes | 118 | 120 | 1.98(1.43-2.76) | 2.19(1.49-3.23) | **0.000\*\*** |
|  | No | 133 | 269 | 1 | 1 |  |
| Attending nightclubs | Yes | 124 | 114 | 2.17(1.56-3.02) | 2.04(1.41-2.96) | **0.000\*\*** |
|  | No | 134 | 268 | 1 | 1 |  |
| SRH Discussion with family/peers | No | 120 | 118 | 1.60(1.16-2.21) | 2.09(1.17-3.75) | **0.013\*** |
|  | Yes | 156 | 246 | 1 | 1 |  |
| Attitude towards SRH issues | Unfavorable | 91 | 147 | 0.638(0.46-0.88) | 0.69 (0.47-1.01) | 0.06 |
|  | Favorable | 198 | 204 | 1 | 1 |  |

\*Significance at $p\text{-value} < 0.05$

## Discussion

This study assessed the prevalence of risky sexual practices and associated factors among taxi and three-wheeled vehicle drivers in Arba Minch Town, Southern Ethiopia. The findings indicate that a substantial proportion of drivers engage in behaviors considered risky for sexual and reproductive health, including multiple sexual partnerships, inconsistent condom use, and sexual intercourse with commercial sex workers.

Overall, the prevalence of lifetime and current risky sexual practices was found to be 46.9% and 37.2%,respectively. The finding is high compared to a study done in Humera, Tigray region (13.7%), in Jimma, Oromia region (42.1%)(3, 4)butless similar to the estimate by a study done in Lalibela, Amhara region (46.5 %)(18),whereas it is lower than studies done in Harrar, Ethiopia, lifetime risky sexual behavior were (50.3 %)(21), South Africa (81%) (22), Turkey (51.9%) (23) and Brazil (61.9%) (24)Bangkok, Thailand (69.5%) (25)and Bangladesh (51%) (26). The observed differences may be explained by variations in study populations, measurement tools for defining risky sexual behavior, socio-cultural contexts, urbanization levels, access to sexual health services, and differences in sample size and study periods. In particular, the occupational mobility of drivers, exposure to urban nightlife environments and varying levels of health promotion interventions across settings may also contribute to the observed heterogeneity.

Living arrangement was significantly associated with risky sexual practices. Drivers living alone were more likely to engage in risky sexual behaviors compared to those living with family members. The finding is supported by studies conducted at BahirDar(5), Dhaka, Bangladesh (26), Poland(13), and the US (1). This might be associated with living alone may reduce social control and increase exposure to peer influence and high-risk social environments. It may also be associated with loneliness and increased engagement in nightlife activities, which can increase opportunities for multiple sexual partnerships and reduce adherence to safer sexual practices.

The absence of discussion on sexual and reproductive health issues with family or friends was also significantly associated with risky sexual practices. Similar findings have been reported in studies from BenishangulGumuz(27) and Mexico(28). However, a study in Ghana demonstrated that family/ friend communication does not affect an individual’s sexual activity(29). This could be due to a lack of perceptions and awareness of the detrimental health consequences of risky sexual practices and a misunderstanding of the importance of condom use. In addition, stigma surrounding sexual health discussions may further limit open communication, thereby increasing vulnerability to unsafe sexual behaviors.

Substance use was another important determinant of risky sexual practices. Drivers who reported substance use had higher odds of engaging in risky sexual behaviors compared to non-users. This finding is consistent with studies from MadaWalabu(30),Bahir-Dar (17), Uganda (31), South Africa(22), and studies in nine Asian Countries (32). Usually, substance use and risky sexual practices go hand in hand(32). This might be due to substance use, which can adversely affect or reduce self-control and increase impulsivity; following substance use also impairs judgment and decision-making. (32). This could also be because substance use creates a favorable opportunity (multiple sexual partners, attending nightclubs, and watching pornography) to improve risky sexual practices of the individuals and lower risk perception and increase willingness to engage in unprotected sex.

Similarly, attending nightclubs was significantly associated with risky sexual practices. Drivers who had a history of nightclub attendance were more likely to engage in risky sexual behaviors compared to those who did not. This is consistent with evidence from Finoteselam(20), South Africa(22), Bangkok, Thailand (26), and Poland (13). This might be nightclubs may provide opportunities for alcohol and substance use, reduced inhibitions, and increased interaction with commercial sex workers, all of which may contribute to unsafe sexual practices and increased risk of sexually transmitted infections and unintended pregnancies.

Exposure to pornographic materials was also significantly associated with risky sexual practices. Drivers who reported watching pornography had higher odds of engaging in risky sexual behaviors. This finding aligns with studies from Bahir-Dar (17), in Tanzania (33)and in the US (1). The reason might be exposure to pornography may shape sexual attitudes and expectations, potentially normalizing multiple partnerships and unprotected sexual practices. It may also increase sexual desire and risk-taking behavior, particularly in contexts where sexual health education and condom use promotion are inadequate.

### Limitations of the study

This study has several limitations. First, the cross-sectional design limits the ability to establish temporal or causal relationships between independent variables and risky sexual practices. Second, the absence of female taxi and three-wheeled vehicle drivers in the sample limits the generalizability of the findings and precludes gender-based comparisons. In addition, self-reported data may be subject to social desirability bias, particularly for sensitive behaviors such as sexual practices and substance use.

## Conclusion

The findings of this study demonstrate that taxi and three-wheeled vehicle drivers in Arba Minch engage in a substantial level of risky sexual practices, representing an important and under-recognized public health concern within this occupational group. Nearly half of the respondents reported lifetime engagement in risky sexual behaviors, while more than one-third reported current involvement.

Multivariable analysis indicated that risky sexual practices were significantly associated with modifiable behavioral and lifestyle factors, including living alone, lack of sexual and reproductive health (SRH) discussions, substance use, exposure to pornography, and attendance at nightclubs. In contrast, socio-demographic and background variables such as age, educational status, vehicle ownership, peer influence in sexual initiation, history of sexually transmitted diseases, and SRH attitudes were not significantly associated in the final model.

These findings highlight that risky sexual behaviors among drivers are largely driven by modifiable and context-specific factors rather than fixed socio-demographic characteristics. This underscores the importance of targeted, behavior-focused interventions.

Accordingly, there is a clear need for multi-sectoral and innovative intervention strategies that extend beyond traditional health service delivery. Integrating SRH education, risk-reduction messaging, and substance use prevention components into driver licensing procedures and periodic refresher training programs could provide a practical and scalable approach to reach this mobile population. Collaboration between transport authorities and public health institutions is essential to institutionalize such interventions.

In addition, strengthening community-based SRH communication and embedding health promotion activities within transport-related settings may contribute to reducing risky sexual behaviors and associated health risks among taxi and three-wheeled vehicle drivers.

## Data Availability

Availability of data and materials: The data supporting the findings of this study are presented within this manuscript. Additional de-identified datasets are available from the corresponding author upon reasonable request.

## Acknowledgments

We would like to express our sincere gratitude to Arba Minch University for its approval and support of this study. We also thank the town health administration for their valuable assistance in facilitating the study and coordinating data collection activities.

We are deeply grateful to all study participants for their time and willingness to take part in this research. Finally, we extend our heartfelt appreciation to the data collectors and supervisors for their dedication and commitment throughout the study.

## Availability of data and materials

The data supporting the findings of this study are presented within this manuscript. Additional de-identified datasets are available from the corresponding author upon reasonable request.

## Authors’ contribution

All authors contributed significantly to this work, including its conception and design, data collection, analysis, and interpretation. They participated in drafting the manuscript and revising it critically for important intellectual content. All authors have approved the final version for publication, agreed on the target journal, and take full responsibility for the integrity and accuracy of all aspects of the study.

